# Risk models based on non-cognitive measures may identify presymptomatic Alzheimer’s disease

**DOI:** 10.1101/2022.03.30.22273204

**Authors:** Jingjing Yang, Shahram Oveisgharan, Xizhu Liu, Robert S Wilson, David A Bennett, Aron S Buchman

## Abstract

**Background:** Alzheimer’s disease is a progressive disorder without a cure. Developing risk prediction models for detecting presymptomatic Alzheimer’s disease using non-cognitive measures is necessary to enable early interventions.

**Objective:** Examine if non-cognitive metrics alone can be used to construct risk models to identify adults at risk for Alzheimer’s dementia and cognitive impairment.

**Methods:** Clinical data from older adults without dementia from the Memory and Aging Project (MAP, n=1179) and Religious Orders Study (ROS, n=1103) were analyzed using Cox proportional hazard models to develop risk prediction models for Alzheimer’s dementia and cognitive impairment. Models using only non-cognitive covariates were compared to models that added cognitive covariates. All models were trained in MAP, tested in ROS, and evaluated by the AUC of ROC curve.

**Results:** Models based on non-cognitive covariates alone achieved AUC (0.800,0.785) for predicting Alzheimer’s dementia (3,5) years from baseline. Including additional cognitive covariates improved AUC to (0.916,0.881). A model with a single covariate of composite cognition score achieved AUC (0.905,0.863). Models based on non-cognitive covariates alone achieved AUC (0.717,0.714) for predicting cognitive impairment (3,5) years from baseline. Including additional cognitive covariates improved AUC to (0.783,0.770). A model with a single covariate of composite cognition score achieved AUC (0.754,0.730).

**Conclusion:** Risk models based on non-cognitive metrics predict both Alzheimer’s dementia and cognitive impairment. However, non-cognitive covariates do not provide incremental predictivity for models that include cognitive metrics in predicting Alzheimer’s dementia, but do in models predicting cognitive impairment. Further improved risk prediction models for cognitive impairment are needed.

## INTRODUCTION

Alzheimer’s disease is a progressive disorder that develops over years. An asymptomatic stage during which Alzheimer’s disease pathology (AD) accumulates is followed by mild cognitive impairment (MCI), while dementia occurs later [1-3]. Alzheimer’s disease is heterogeneous; most demented individuals transition through all three clinical stages i.e., from no cognitive impairment (NCI) to MCI followed by dementia. Some individual’s transition directly from NCI to dementia. On the other hand, some never progress beyond MCI and 1/3 with NCI prior to death also show pathologic AD at death.

Early detection of adults with normal cognition at risk for Alzheimer’s disease cognitive traits is crucial for early targeted treatments [4, 5]. This underscores the widespread efforts to develop risk prediction models for detecting presymptomatic Alzheimer’s disease. Yet, using cognitive test scores as a predictor for the incident cognitive impairment (MCI or Alzheimer’s dementia) may be statistically problematic as the same test scores may be used as model predictors as well as for the diagnostic characterization of cognitive status, which is the outcome the model seeks to identify. Moreover, comprehensive cognitive testing requires in-person testing that is not widely available. This highlights the potential benefits of identifying non-cognitive covariates that may be obtained remotely and contain predictive information to identify cognitively normal adults at risk for incident cognitive impairment and Alzheimer’s dementia.

Alzheimer’s disease is a complex disorder that not only negatively affects cognition, but also adversely affects other important non-cognitive aging phenotypes [6]. For example, accumulation of AD is associated with body mass index (BMI) decline or impaired motor function that may precede and predict incident cognitive impairment and Alzheimer’s dementia [7-11]. Gait velocity and variability are associated with cognitive decline and gait is shown as a proxy for overall health and as an index of cognitive decline [12]. These data suggest that developing a risk profile based on non-cognitive covariates may facilitate the identification of at risk adults during the earliest stages of Alzheimer’s disease.

To test this hypothesis, it is crucial to examine non-cognitive and cognitive covariates alone and together to compare model performance and identify the optimal covariates for predicting incident Alzheimer’s dementia and incident cognitive impairment. This study used clinical data from older adults without dementia participating in one of two prospective community-based cohort studies of aging, Rush Memory and Aging Project (MAP) and the Religious Orders Study (ROS) [13]. We trained models in MAP and validated model prediction performance in ROS.

## MATERIALS AND METHODS

### Participants

Participants were community dwelling older persons enrolled in one of two ongoing cohort studies of aging and dementia, MAP (n=1179) and ROS (n=1103). Participants were enrolled without known dementia; 1742 with NCI and 540 with MCI at enrollment (**Tables S1-S2**). All agreed to annual clinical evaluations and autopsy at the time of death. The duration of follow up for participants ranged from 2 to 26 years, with a median of 8-year and standard deviation 5.42 (**Supplementary Figure 1**). Both cohorts employ a harmonized data collection battery administered by the same research assistants facilitating joint analyses.

**Fig 1.**
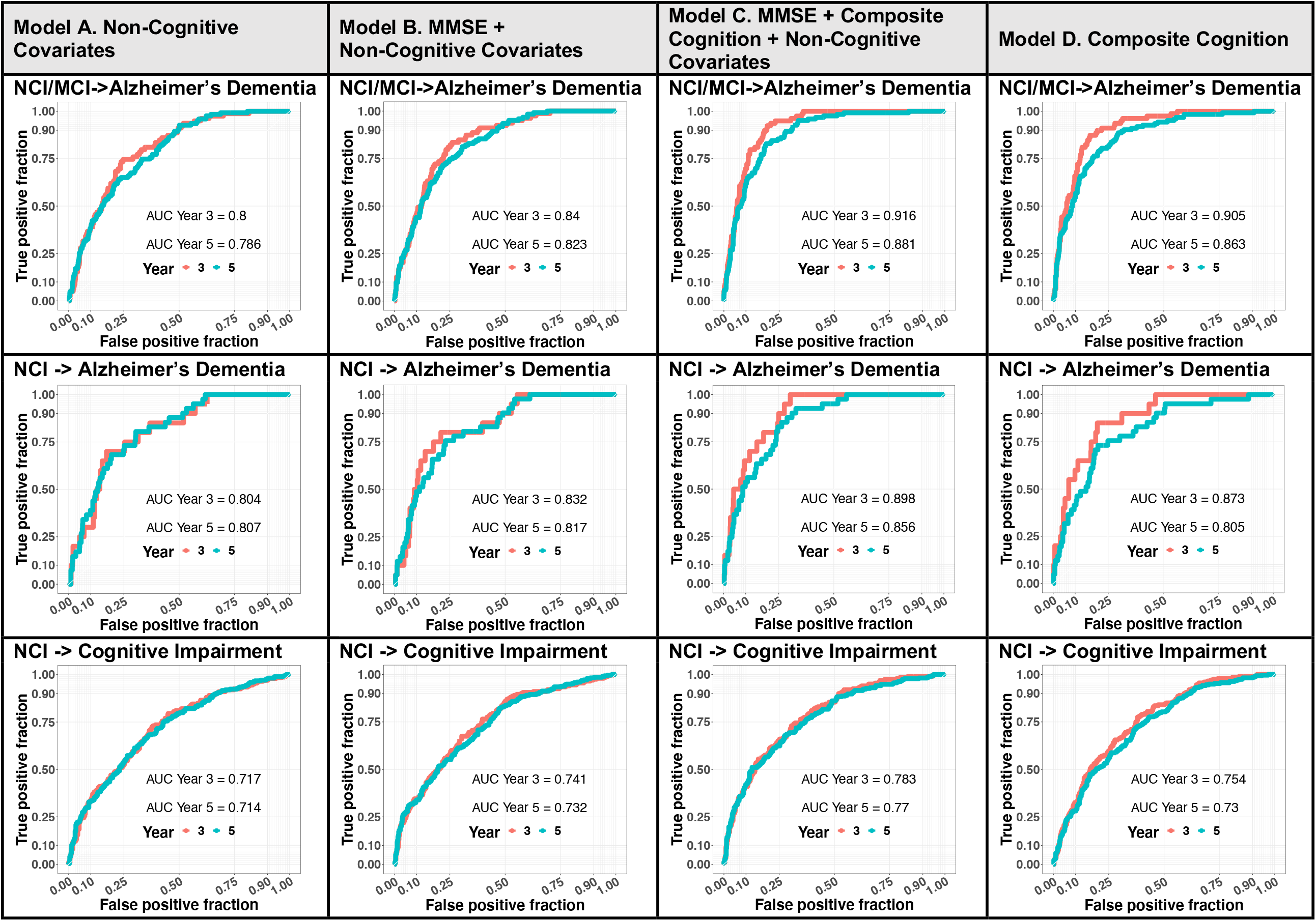
ROC plots of risk prediction for incident Alzheimer’s dementia and incident cognitive impairment with non-cognitive and cognitive covariates alone and together. <u>Model A</u> uses 55 non-cognitive covariates; <u>Model B</u> 55 non-cognitive covariates + MMSE; <u>Model C</u> 55 non-cognitive covariates+ MMSE+ Composite Cognition; <u>Model D</u> uses only Composite Cognition. **ROW 1 No dementia (NCI or MCI) at baseline and incident Alzheimer’s Dementia:** Model B vs A showed significant improvement with p-values=(4.85 × 10^−4^, 1.09 × 10^−4^) for Year (3, 5). Model C vs B showed significant improvement with p-values=(7.55 × 10^−7^, 9.15 × 10^−6^) for Year (3, 5). Model C vs D for Year 3 (p-value = 0.156) but significant improvement for Year 5 (p-value = 0.033). **ROW 2 NCI at baseline and incident Alzheimer’s Dementia:** Model B vs A showed significant improvement only for Year 3 with p-value=0.042 but not for Year 5 with p-value=0.185. Model C vs A/B showed significant improvement p-values=∼(0.004, 0.019) for Year (3, 5). Model C vs D showed comparable performance for Year 3 with p-value=0.156 but significant improvement for Year 5 with p-value=0.025. **ROW 3 NCI and incident cognitive impairment:** Model B vs A showed significant improvement p-values p-values=(0.002, 0.008) for Year (3, 5). Model C vs B showed significant improvement p-values=(0.001, 0.001) for Year (3, 5). Model C vs D showed significant improvement p-values=(0.029, 0.002) for Year (3, 5).

### Cognitive Assessment and Diagnoses

Detailed structured clinical examinations were administered annually to participants in both studies. The neuropsychological battery includes 17 tests that assess five domains of cognitive ability. Raw test scores were standardized per test using baseline means and standard deviations (SDs) of both cohorts; the resulting Z-cores were then averaged across the 17 cognitive tests to derive a summary composite cognition score and scores for five cognitive abilities including: episodic memory, semantic memory, working memory, visuospatial abilities, and perceptual speed.

A neuropsychologist and a neurologist with expertise in dementia reviewed annual cognitive testing and classified cognitive status of participants as NCI, MCI, Alzheimer’s dementia, or other dementias according to established NIA-AA criteria [13-16]. We excluded individuals who developed non-Alzheimer’s dementia from these analyses.

This study first examined models which predicted transition from no dementia (NCI or MCI) to Alzheimer’s dementia (**Fig 1, Row 1**). Second, we examined models which predicted transition from NCI to Alzheimer’s dementia (**Fig 1, Row 2**). Third, we studied the outcome of incident cognitive impairment which was defined by the first annual assessment when an individual who was previously cognitively normal (NCI) transitioned to cognitive impairment manifested as either MCI or Alzheimer’s dementia (**Fig 1, Row 3**).

### Other Clinical Covariates

Five groups of clinical variables (a total of 57) were considered potential covariates in the Cox proportional hazard regression models for risk prediction (**Table 1; Tables S3-S4**) –– i) *Common Alzheimer’s disease risk factors* including age, sex, education, Mini Mental State Exam (MMSE) score [17] and *APOE* E4 allele; ii) *Health measures* such as blood pressure, depression, and cardiovascular diseases; iii) *Medication usage* such as lipid lowering medications, and antidepressants; iv) *Variables uniquely profiled by MAP/ROS* such as composite cognition scores based on 17 cognitive tests, self-reported physical activity and social network size [13]; v) *Motor and sleep metrics* such as dexterity, hand strength, gait function, four parkinsonian signs [16], and four self-reported variables about sleep quality and duration [15].

**Table 1.** Select clinical characteristics of the analytic cohort at baseline (n=1179 in MAP; n=1103 in ROS).

| Group | Variable | Mean (SD) or N (%) |
| --- | --- | --- |
| **Demographics and Common Alzheimer's Risk Factors** | Mild cognitive impairment (N) | 541 (23.7) |
|  | Follow-up years (2-26) | 8 (5.42) |
|  | Age (years, 53-102) | 77.4 (7.51) |
|  | Sex (males, N) | 577 (25.2) |
|  | Education (years, 0-30) | 16.5 (3.65) |
|  | BMI (9.09, 62.91) | 27.52 (5.50) |
|  | MMSE Score (18-30) | 28.37 (1.73) |
|  | APOE E2 (carriers, N) | 330 (14.4) |
|  | APOE E4 (carriers, N) | 513 (22.4) |
| **Covariates Uniquely Profiled by ROS/MAP** | Composite cognition test Z-score (-1.95, 1.47) | 0.32 (0.51) |
|  | Physical activity (0-35) | 3.19 (3.79) |
|  | Social network size (0, 162) | 7.58 (7.52) |
|  | Maternal education (years, 0-20) | 9.66 (3.79) |
|  | Paternal education (years, 0-24) | 9.60 (4.26) |
|  | Anxiety (0-10) | 1.28 (1.69) |
|  | Proneness to psychological distress (0-42) | 15.69 (6.44) |
|  | Tendency to be social, active, and optimistic (5, 24) | 15.52 (3.14) |
|  | Early life socioeconomic status Z-score (-2.72, 2.15) | 0.02 (0.74) |
|  | Anger (0-10) | 1.76 (1.88) |
|  | TOMM40_S (N) | 1,126 (49.2) |
|  | TOMM40_L (N) | 371 (16.2) |
|  | TOMM40_VL (N) | 1,114 (48.7) |
| **Motor Metrics** | Dexterity (0, 1.85) | 1.02 (0.16) |
|  | Gait function (0, 2.13) | 1.05 (0.27) |
|  | Hand strength (0, 2.36) | 1.07 (0.30) |
|  | Bradykinesia (0, 85) | 8.82 (10.6) |
|  | Parkinsonian gait (0, 85.18) | 12.16 (13.45) |
|  | Rigidity (0, 60) | 2.15 (5.88) |
|  | Tremor (0, 69.69) | 2.23 (5.02) |
| **Self-reported Sleep Metrics** | Sleep latency (1, 9) | 3.63 (1.14) |
|  | Sleep consolidation (1, 8) | 2.92 (1.31) |
|  | Daytime sleepiness (1, 5) | 3.21 (1.33) |
|  | Sleep quality (1, 5) | 2.10 (1.05) |

These 57 clinical covariates were selected by excluding variables with a high proportion of missing values (>20%) and those that were highly correlated with other clinical variables (correlation>0.95). Missing baseline values of longitudinal variables were imputed from the corresponding values at participant’s nearest past visit. A total of 345 individuals with missing cross-sectional variables were excluded, leaving n=1179 MAP and n=1103 ROS participants for these analyses. A heat map showing the correlations of the covariates examined in these models is shown in **Supplementary Figure 2**.

**Fig 2.**
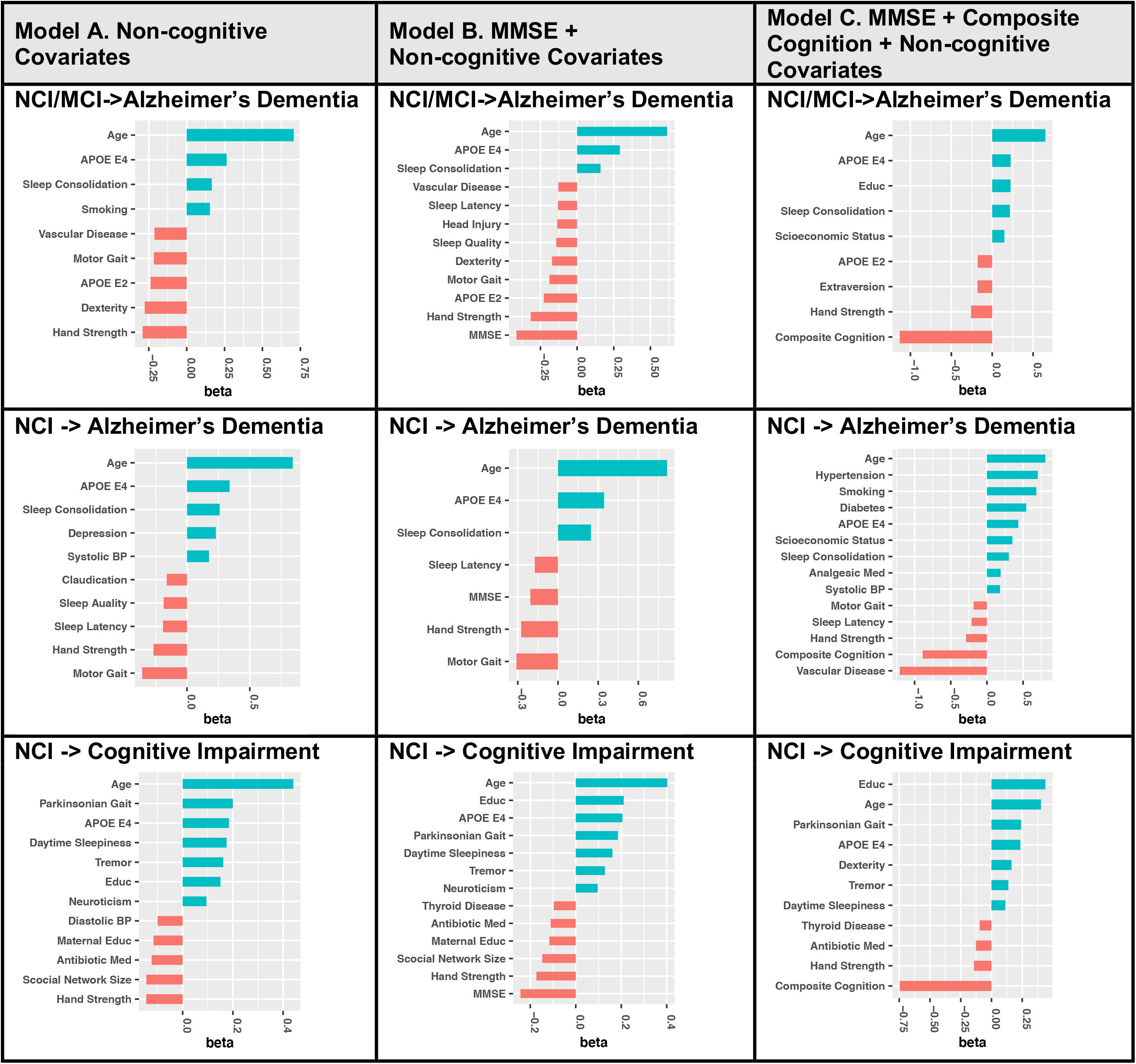
Important predictive covariates predicting incident Alzheimer’s dementia and incident cognitive impairment. **ROW 1 No dementia (NCI or MCI) at baseline and incident Alzheimer’s Dementia:** Age and APOE E4 allele were identified as the top positive risk factors in all three models. Hand strength was the top protective factor in Model A when no cognitive covariates were considered. MMSE was the top protective factor in Model B when composite cognition was not considered. Composite cognition score was the top protective factor in Model C where all clinical covariates were considered. **ROW 2 NCI at baseline and incident Alzheimer’s Dementia:** Age was identified as the top positive risk factor in all three models. Motor gait and hand strength were top protective factor in Model A and Model B when composite cognition score was not considered. Instead of composite cognition score, vascular disease history at baseline was the top “protective” factor in Model C, which might lead to death before the development of Alzheimer’s dementia. **ROW 3 NCI and incident cognitive impairment:** Age and education were identified as top positive risk factors in all three models, where education years were positively correlated with the number of follow-up years in ROS/MAP. Hand strength and social network size were top protective factor in Model A when no cognitive covariates were considered. MMSE was the top protective factor in Model B when composite cognition was not considered. Composite cognition score was the top protective factor in Model C where all clinical covariates were considered.

### Analytic Approach

The annual cognitive status diagnosis and the follow up year were used to identify the first occurrence of Alzheimer’s dementia or incident cognitive impairment. For each participant, the year of enrollment is considered as baseline (time 0), the year of first diagnosis of Alzheimer’s dementia and cognitive impairment (MCI or Alzheimer’s dementia) is considered as the time when the event occurs (incident Alzheimer’s dementia or incident cognitive impairment), and the last visit of participants without the considered event during all follow-ups is considered as the right censored time for living participants or the time of death for dead participants. Less than 10% of participants developing Alzheimer’s dementia progressed directly from NCI to Alzheimer’s dementia. Sample size distributions with respect to cognitive status at baseline and the cognition event types are included in **Tables S1-S2**.

We fit separate Cox proportional hazard risk prediction models [18-21] for incident Alzheimer’s dementia or incident cognitive impairment while accounting for the competing risk of death. In the first stage of our analysis, we examined the prediction accuracy of using non-cognitive covariates alone in the Cox models (Model A). Then, we examined if adding cognitive covariates improved the model accuracy assessed. Model performance, i.e., discrimination was evaluated by the prediction area under curve (AUC) values of receiver operating characteristic curve (ROC) [22] for the risk of developing Alzheimer’s dementia and cognitive impairment 3, 5 years from the analytic baseline. AUC is equal to the probability that a classifier will rank a randomly chosen individual with the outcome of interest (event or case) higher than a randomly chosen individual without impaired cognition (control), assuming ‘case’ ranks higher than ‘control’ [22]. That is, the derived risk prediction model with higher AUC is expected to predict the risk of developing Alzheimer’s dementia or cognition impairment with higher accuracy. ‘control’ [22]. That is, the derived risk prediction model with higher AUC is expected to predict the risk of developing Alzheimer’s dementia or cognition impairment with higher accuracy.

Next, we added the MMSE score (Model B) and then repeated the model a third time adding the composite cognition score (Model C). In further analyses, we sought to isolate the prediction accuracy of different combinations of cognitive metrics alone including: composite cognition score (Model D), MMSE score (Model E), as well as the five cognitive ability scores (Model F) used for its construction and the 17 individual cognitive test scores (Model G). Since all covariates were standardized, the magnitudes of covariate coefficients in the Cox models are comparable with respect to predictivity.

By selecting a risk score threshold corresponding the highest overall classification accuracy (the proportion of correct discriminations in test ROS samples), we compared the overall classification, sensitivity (the proportion of true positive predictions in all test cases, i.e., true positive fraction), and specificity (the proportion of true negative predictions in all test controls, i.e., 1 – false positive fraction), for Models A-D. Samples with risk scores greater than the selected threshold were predicted as positives or otherwise negatives. The sensitivity and specificity corresponding to a selected risk score threshold reflect risk model performance at one point in the ROC curves.

Backward variable selection was implemented during the training of Cox models that consider more than one covariate. In particular, we started with the full Cox model with all potential covariates; proceeded with a temporary Cox model by excluding the covariate with the least significant p-value; compared their C-statistics (equivalent to AUC) and conducted ANOVA test to examine if the temporary Cox model would be improved over the current Cox model; accepted the temporary Cox model if the C-statistic was improved in the temporary Cox model or otherwise the ANOVA test p-value<0.05. We continued to exclude the covariate with the least significant p-value from the accepted Cox model until the proposed temporary Cox model was not accepted or when there was only one covariate remained in the model.

For Alzheimer’s dementia, we first considered all individuals including adults with either NCI or MCI at baseline, and then only individuals with NCI at baseline. Modeling incident cognitive impairment considered only individuals with NCI at baseline. All Cox models were trained using data from MAP participants and validated (i.e., tested) with data from ROS participants [23]. We tested the respective AUC improvement in 3, 5 years of two models by a nonparametric test that accounts for the correlated nature of the ROC curves [24]. We also evaluated the prediction accuracy, sensitivity, and specificity by choosing the prediction risk score threshold with respect to the highest overall accuracy.

### Association Tests between Predicted Risk Scores of Alzheimer’s Dementia and Brain Pathologies

We investigated the association between predicted risk scores (by the full Model C) of NCI/MCI samples developing Alzheimer’s dementia in 3, 5 years in our test ROS cohort and Alzheimer’s disease related dementia (ADRD) neurodegenerative and cerebrovascular disease brain pathologies –– Amyloid, Tangles, Global AD pathology (Gpath), NIA Reagan pathology (NIA), Lewy body pathology (LewyBody), Hippocampal Sclerosis (Hippo), TDP-43, Macroinfarct (Macro), Microinfarct (Micro), Atherosclerosis (Arthero), Arteriolosclerosis (Arteriol), Cerebral amyloid angiopathy (CAA). Correlation tests were conducted for quantitative pathologies (Amyloid, Tangles, and Gpath), and two-sample t-tests were conducted for the remain dichotomized pathologies (present vs. not present). Only deceased ROS participants who underwent autopsy and had profiled pathologies were tested (∼50%).

### Standard Protocol Approvals, Registrations, and Patient Consents

Both MAP and ROS studies were approved by an Institutional Review Board of Rush University Medical Center. Written informed consent was obtained from all study participants as was an Anatomical Gift Act for organ donation.

### Data Availability

All data analyzed in this study are de-identified and available to any qualified investigator with application through the Rush Alzheimer’s Disease Center Research Resource Sharing Hub, https://www.radc.rush.edu, which has descriptions of the studies and available data.

## RESULTS

### Risk Models Predicting Alzheimer’s Dementia

#### Overall model performance

Cox proportional hazard model trained using only non-cognitive covariates showed good model performance [25] for predicting Alzheimer’s dementia 3, 5 years from baseline (**Fig 1, Row 1 Model A**). Stepwise addition of the cognitive covariate MMSE and then composite cognition yielded further significant improvement in the initial model performance of using only non-cognitive covariates (**Fig 1, Row 1 Model B & Model C)**. The performance accuracy of the full model including non-cognitive covariates, MMSE and composite cognition score was similar to the performance of a model including composite cognition score alone (Model D) for Year 3. The full model outperformed Model D for Year 5 (**Fig 1, Row 1 Model C & Model D**).

#### Important clinical predictors

As expected, composite cognition score had the largest protective effect size when it was considered in the full model (**Fig 2, Row 1 Model C**). Similarly, in the model including non-cognitive and MMSE covariates (**Supplementary Table 5**), we observed that MMSE had the largest protective effect size, followed by hand strength and *APOE* E2. In contrast, increasing age and the presence of *APOE* E4 had the largest effect size for increased risk of Alzheimer’s dementia, when only non-cognitive covariates were considered in Model A.

#### Cognitive metrics as predictors

To better understand the role of cognitive measures in predicting Alzheimer’s dementia, we compared the model performance of different combinations of cognitive metrics alone for predicting Alzheimer’s dementia (**Supplementary Figure 3 Row 1)**. Model performance for MMSE predicting Alzheimer’s dementia (**Supplementary Figure 3, Row 1 Model E**) was comparable with a model based only on non-cognitive measures (**Fig 1, Row 1 Model A**). The various cognitive metrics based on 17 cognitive tests alone (**Supplementary Figure 3, Row 1 Model F and Model G**) showed similar excellent model performance that was not improved by the addition of the non-cognitive covariates or MMSE. Of all the cognitive indices, episodic memory had the largest protective effect size (**Supplementary Figure 4, Row 1**).

#### Exclude baseline MCI patients

Analyses of individuals without dementia include adults with NCI or MCI at baseline. In further analyses, we restricted our analyses only to individuals with baseline NCI to see if the same trends were observed despite the truncated heterogeneity of cognitive function by exclusion of individuals with baseline MCI. Model performance for non-cognitive covariates for adults with baseline NCI alone was similar to models that also included adults with baseline MCI (**Fig 1, Row 2 Model A versus Row 1 Model A**). There was improved model performance when cognitive covariates MMSE or composite cognition scores were added to the non-cognitive covariates (**Fig 1, Row 2 Model B and Model C**). Yet, improvement in model performance for predicting dementia in adults with baseline NCI alone was less marked compared to models that consider adults with baseline NCI or MCI (**Fig 1, Row 2 vs. Row 1, Model B and Model C**).

#### Classification accuracy

By comparing the prediction accuracy, sensitivity, and specificity corresponding to a selected risk score threshold ensures the highest overall accuracy per model for Models A-D. The pattern in **Table 2** was similar to **Fig 1, Row 1 and Row 2**. That is, the full Model C performed the best with overall accuracy for Year (3, 5), that outperformed Model A and Model B for Year (3, 5) when composite cognition was not considered, and outperformed Model D for Year 5 when only composite cognition was considered. The full Model C performed comparably with Model D for Year 3.

**Table 2.** Prediction accuracy (with 95% confidence interval), sensitivity, and specificity with respect to selected risk score thresholds that ensure the highest overall accuracy. Values in this table are reflecting risk model prediction performance at one point in the ROC curves as shown in Fig1, with corresponding risk score thresholds. Samples with predicted risk scores greater than the selected threshold were considered as Predicted Positives (incident Alzheimer’s dementia or incident cognitive impairment), otherwise Predicted Negatives (not developing Alzheimer’s dementia or NCI).

|  |  | Model A.<br>Non-Cognitive<br>Covariates |  | Model B.<br>MMSE +<br>Non-Cognitive<br>Covariates |  | Model C.<br>MMSE +<br>Composite<br>Cognition + Non-Cognitive<br>Covariates |  | Model D.<br>Composite<br>Cognition |  |
| --- | --- | --- | --- | --- | --- | --- | --- | --- | --- |
|  |  | Y3 | Y5 | Y3 | Y5 | Y3 | Y5 | Y3 | Y5 |
| **NCI/MCI ->**<br>**Alzheimer's**<br>**Dementia** | **Accuracy<sup>a</sup>**<br>**(95% CI)** | **0.664**<br>**(0.637,**<br>**0.694)** | **0.616**<br>**(0.586,**<br>**0.644)** | **0.761**<br>**(0.735,**<br>**0.786)** | **0.707**<br>**(0.679,**<br>**0.734)** | **0.856**<br>**(0.834,**<br>**0.876)** | **0.816**<br>**(0.792,**<br>**0.838)** | **0.865**<br>**(0.844,**<br>**0.885)** | **0.784**<br>**(0.758,**<br>**0.808)** |
|  | **Sensitivity<sup>b</sup>** | **0.810** | **0.804** | **0.810** | **0.804** | **0.810** | **0.804** | **0.810** | **0.804** |
|  | **Specificity<sup>c</sup>** | **0.655** | **0.591** | **0.757** | **0.694** | **0.860** | **0.817** | **0.870** | **0.781** |
| **NCI ->**<br>**Alzheimer's**<br>**Dementia** | **Accuracy<sup>a</sup>**<br>**(95% CI)** | **0.688**<br>**(0.655,**<br>**0.719)** | **0.701**<br>**(0.669,**<br>**0.732)** | **0.790**<br>**(0.761,**<br>**0.818)** | **0.695**<br>**(0.662,**<br>**0.726)** | **0.818**<br>**(0.790,**<br>**0.844)** | **0.763**<br>**(0.733,**<br>**0.791)** | **0.806**<br>**(0.778,**<br>**0.832)** | **0.645**<br>**(0.611,**<br>**0.677)** |
|  | **Sensitivity<sup>b</sup>** | **0.800** | **0.804** | **0.800** | **0.804** | **0.800** | **0.804** | **0.800** | **0.804** |
|  | **Specificity<sup>c</sup>** | **0.685** | **0.695** | **0.790** | **0.689** | **0.818** | **0.761** | **0.806** | **0.636** |
| **NCI ->**<br>**Cognitive**<br>**Impairment** | **Accuracy<sup>a</sup>**<br>**(95% CI)** | **0.650**<br>**(0.617,**<br>**0.682)** | **0.635**<br>**(0.601,**<br>**0.667)** | **0.662**<br>**(0.629,**<br>**0.693)** | **0.648**<br>**(0.614,**<br>**0.680)** | **0.701**<br>**(0.669,**<br>**0.732)** | **0.685**<br>**(0.679,**<br>**0.716)** | **0.672**<br>**(0.639,**<br>**0.703)** | **0.657**<br>**(0.624,**<br>**0.689)** |
|  | **Sensitivity<sup>b</sup>** | **0.700** | **0.700** | **0.700** | **0.700** | **0.700** | **0.700** | **0.700** | **0.700** |
|  | **Specificity<sup>c</sup>** | **0.635** | **0.604** | **0.650** | **0.623** | **0.702** | **0.678** | **0.664** | **0.636** |

a. Accuracy= (# True Positive Predictions + # True Negative Predictions) / (# of test samples)
b. Sensitivity = (# True Positive Predictions) / (# Positives in test samples) = True Positive Fraction
c. Specificity = (# True Negative Predictions) / (# Negatives in test samples) = 1 - False Positive Fraction

### Association between Predicted Risk Scores for Alzheimer’s Dementia and Mixed-Brain Pathologies

Accumulating evidence suggests that clinical Alzheimer’s dementia is most commonly associated with mixed brain pathologies. We examined the association between predicted risk scores of Alzheimer’s dementia by the full risk model for NCI/MCI samples (**Fig 1, Row 1, Model C**) and 12 different postmortem indices of ADRD pathologies in our test ROS cohort. Using the significance threshold with Bonferroni correction for testing 12 pathologies (-log10(0.05/12)), we found that the risk scores predicted by the full Model C for developing Alzheimer’s dementia in 3, 5 years were associated with 5 out of 12 ADRD pathologies, including both neurodegenerative pathologies (Tangles, global AD pathology, NIA Reagan) and cerebrovascular disease pathologies (Atherosclerosis, Cerebral amyloid angiopathy). These results (**Supplemental Figure 5**) showed that the risk scores predicted by our derived models were significantly associated with mixed-brain pathologies.

### Risk Models Predicting Cognitive Impairment

#### Overall model performance

Cox proportional hazard model trained using only non-cognitive covariates showed adequate model performance [25] for predicting cognitive impairment 3 and 5 years from baseline (**Fig 1, Row 3 Model A**). Yet, discrimination for predicting cognitive impairment was lower than the model predictions of Alzheimer’s dementia (**Fig 1, Row 3 vs. Row 1 Model A**). Stepwise addition of MMSE and then composite cognition yielded further improvement in the initial model performance (**Fig 1, Row 3 Model B and Model C**). AUC for all models predicting cognitive impairment were lower than models predicting Alzheimer’s dementia (**Fig 1 and Supplementary Figure 3**). The performance accuracy of the full model that included non-cognitive, MMSE and composite cognition score showed better model performance compared to models with the composite cognition alone (**Fig 1, Row 3 Model C vs. Model D**).

#### Important clinical predictors

By examining the effect sizes in the Cox model using non-cognitive and MMSE covariates (**Fig2; Supplementary Table 5**), we observed that MMSE had the largest protective effect size and age had the largest effect size for increased risk of cognitive impairment. Composite cognition still had the largest protective effect for reducing the risk of cognitive impairment when it was considered in the full model. We observed that age and *APOE* E4 were associated with an increased risk of both Alzheimer’s dementia and cognitive impairment. Better motor function was associated with reduced risk of Alzheimer’ dementia (hand strength and gait) and cognitive impairment (hand strength and parkinsonism) (**Fig 2; Supplementary Table 5**).

#### Cognitive function metrics

Next, we compared model performance for several cognitive metrics alone for predicting cognitive impairment. Model performance for MMSE alone (**Supplementary Figure 3, Row3 Model E**) was poor and significantly lower than a model based on all the non-cognitive covariates together (**Fig 1, Row 3 Model A**). Models using scores of five cognitive abilities (**Supplementary Figure 3, Row3 Model F**) or using the 17 individual cognitive test scores (**Supplementary Figure 3, Row3 Model G**) had comparable prediction AUC as the model using composite cognition alone (**Supplementary Figure 3, Row3 Model D**). These results together with the joint modeling of cognitive and non-cognitive covariates (**Fig1, Row 3**) show that non-cognitive clinical covariates contain additional predictive information for predicting cognitive impairment not be accounted for by using cognitive covariates alone.

#### Classification accuracy

By comparing the prediction accuracy, sensitivity, and specificity corresponding to a selected risk score threshold ensures the highest overall accuracy per model for Models A-D (**Table 2**). These models show a similar pattern to **Fig 1, Row 3**. That is, the full Model C performed the best with the highest overall accuracy and specificity for Year (3, 5) performing better than models that did not include global cognition score (A and B) or a model with global cognition score alone (Model D). So, joint models of non-cognitive and cognitive measures provide better predictions of cognitive impairment than cognitive measures alone.

## Discussion

### Risk models for incident Alzheimer’s dementia

This study used a wide range of non-cognitive covariates to construct multifactorial risk models that as hypothesized yielded good model performance for predicting incident Alzheimer’s dementia. An important aspect of this study was that we examined model performance achieved by non-cognitive covariates and cognitive function metrics alone and jointly in adults with no dementia. This analytic approach allows us to isolate the potential improvement in model performance due to non-cognitive or cognitive covariates. Our results highlight the effectiveness of cognitive measures alone in predicting Alzheimer’s dementia.

While risk models like the Framingham Risk Score have been developed to predict vascular events [26], there is no widely accepted risk model for the prediction of Alzheimer’s dementia or cognitive impairment [5, 27-32] (**Supplementary Table 6**). In addition, the aging field has not converged on a parsimonious set of dementia risk factors that must be included when developing risk models for cognitive outcomes. Thus, many studies have included cognitive metrics together with non-cognitive covariates [33, 34] and many other studies have not included cognitive measures [5, 27, 28, 30, 35]. Given the varied covariates employed in prior publications, it is not surprising that model discrimination has ranged from an AUC of 0.55 to 0.95 for predicting Alzheimer’s dementia. However, even studies reporting models with adequate discrimination have not usually replicated these findings in independent cohorts. Moreover, there has not been a systematic effort to clarify the independent roles of non-cognitive versus cognitive metrics for predicting Alzheimer’s dementia as well as consensus about whether to include cognitive measures as both predictors and outcome in the same models. Nonetheless, review of these past studies suggests that overall, model performance for predicting Alzheimer’s dementia in very old adults was lower when risk models were based on non-cognitive metrics alone [28].

Our results, validated in an independent cohort, suggest that risk models using non-cognitive covariates show good model performance and predict Alzheimer’s dementia. We also showed that this good performance was further improved by adding cognitive scores. Yet, further analyses showed that the risk models based on composite cognitive metrics alone achieved better model performance than the model based on non-cognitive covariates and MMSE for Alzheimer’s dementia (**Fig 1, Row 1**). By restricting our modeling only to adults with NCI, we showed that non-cognitive variables add additional predictive information that cannot be accounted for by the composite cognitive scores alone for predicting Alzheimer’s dementia (**Fig 1, Row 2**). These results may have important clinical consequences for public health efforts to identify adults with presymptomatic Alzheimer’s disease, i.e., NCI at risk for future dementia. Further work is needed to identify non-cognitive covariates or AD biomarkers that may improve risk models in adults with NCI at the earliest stages of Alzheimer’s disease.

### Risk models for incident cognitive impairment

Transition from NCI to MCI, an earlier stage in the trajectory of Alzheimer’s disease, occurred in almost all participants (>90%) who developed Alzheimer’s dementia in MAP/ROS studies. Since the biology underlying earlier and later stages of Alzheimer’s disease may vary, their respective risk models may be different [3]. Compared to studying Alzheimer’s dementia, larger sample size and wider range of clinical variables are needed for studying cognitive impairment that includes transition from NCI to either MCI or Alzheimer’s dementia. Thus, there are far fewer studies which have developed risk models for predicting cognitive impairment as compared to dementia and even fewer have validated findings in independent samples. Model performances reported for predicting cognitive impairment have varied widely (**Supplementary Table 7**) [34, 36-38]. We are unaware of studies that isolated the independent and joint contributions of non-cognitive and cognitive metrics for model discrimination of incident cognitive impairment.

Our study found that risk models for cognitive impairment based on only non-cognitive covariates showed adequate model performance but consistently poorer than prediction models for Alzheimer’s dementia (**Fig 1 Row 2 vs Row 3**). While adding cognitive covariates in addition to non-cognitive covariates improved model performance predicting cognitive impairment, overall performance never equaled model performance for predicting Alzheimer’s dementia. These findings highlight the importance of further studies to identify additional clinical metrics or biomarkers to improve the prediction of cognitive impairment.

### Risk models for cognitive impairment versus Alzheimer’s dementia

An additional strength of the current study design is that we compared the performances of models predicting cognitive impairment and Alzheimer’s dementia using the same covariates and individuals with NCI at baseline. In addition, we validated our results in an independent sample of older adults. Model performance for predicting Alzheimer’s dementia was much better than models predicting cognitive impairment. These findings may derive from the differences in the symptoms and diagnoses of MCI versus Alzheimer’s dementia. That is, discriminating symptoms of Alzheimer’s dementia from NCI/MCI is generally much easier than discriminating symptoms of MCI from NCI. There is more random error and instability in the diagnosis of MCI compared to dementia. Many prior studies have reported lower sensitivity and specificity for cognitive instruments used to identify MCI compared to Alzheimer’s dementia [39]. Individuals with MCI show more fluctuations between NCI and MCI during repeated cognitive testing. In contrast, once individuals receive a diagnosis of dementia, they are less likely to revert back to NCI/MCI on repeated testing.

Yet, varied model performance for predicting cognitive impairment and Alzheimer’s dementia may inform differences in the respective biology underlying these Alzheimer’s clinical traits [40]. Additionally, the associations between a specific risk factor with dementia may vary over time. For example, high BMI in mid-life may be related to an increased risk of dementia, while high BMI in much older adults may be related to decreased risk of dementia [41]. Analogously, in earlier stages of Alzheimer’s disease, the burden and location of AD may lead to impaired non-cognitive function like motor function to a greater extent than cognition. In contrast, during its later stages the widespread accumulation of AD throughout cognitive brain regions may have a larger pervasive negative impact on cognition. While cognitive and motor decline in older adult are both commonly associated with mixed-brain pathologies, different combinations of pathologies may be associated with cognitive and motor phenotypes in the same individual [6, 11]. This might account for better performance achieved by cognitive metrics for Alzheimer’s dementia and the lack of incremental validity of non-cognitive covariates when considered together with cognitive metrics for Alzheimer’s dementia. This might also account for the incremental predictivity of non-cognitive covariates in addition to cognitive metrics for incident cognitive impairment.

### Implications of the current findings

The reconceptualization of Alzheimer’s disease as a chronic progressive disorder that develops over many years suggests that treatments during its earliest stage when adults manifest normal cognitive function may offer the best chance of preventing future cognitive impairment. However, this opportunity requires the identification of at-risk older adults who are likely to benefit from early interventions. Our current findings lend further support for accumulating evidence that Alzheimer’s disease affects diverse cognitive and other non-cognitive aging phenotypes such as physical frailty and sarcopenia [6, 10, 12, 42]. Thus, our findings are timely as they inform on how to optimize the construction of models to identify at risk older adults by including a wider array of motor, cognitive and behavioral metrics [1, 2, 43, 44]. To date there is no consensus about whether cognitive function metrics can or should be included together with non-cognitive function metrics for predicting Alzheimer’s dementia. The current results demonstrate the importance of addressing this issue since including cognitive function metrics can impact model performance and the value added for non-cognitive measures used to construct risk models for different Alzheimer’s cognitive phenotypes.

Examining important predictive covariates selected in our Cox risk models highlights the importance of motor function (e.g., hand strength and Parkinsonian gait score) as one of the most important non-cognitive covariates in predicting both incident Alzheimer’s dementia and incident cognitive impairment. Although our previous study showed that when cognitive metrics are considered motor function did not show incremental predictivity for Alzheimer’s dementia [45], this study showed that motor metrics provide additional predictive information for both Alzheimer’s dementia and cognitive impairment (**Fig 2; Supplementary Table 5**). Motor planning and ongoing attention is necessary for all movement. Thus, the fact that “we must think first and then move next” may explain why motor metrics predict cognitive impairment. Yet conventional motor testing only quantifies movement and does not assess specific cognitive indices underlying motor planning or executive and attentional resources crucial for movement execution. Instrumented gait testing with unobtrusive sensors can be used to quantify cognitive mobility metrics which precede movement [46, 47]. Further work is needed to determine if adding additional cognitive mobility metrics may improve predictions of adults at risk for Alzheimer’s dementia and cognitive impairment.

Also, broadening the motor metrics used to predict cognitive impairment might help circumvent statistical concerns about confounding predictors with model outcomes as cognitive mobility metrics are not part of current cognitive batteries used to document dementia or cognitive impairment. While there are many knowledge gaps in our understanding of the trajectory for the accumulation of AD pathology, the time course for β-amyloid and tau-tangles during the early and later stages of Alzheimer’s disease may vary; so fluid biomarkers of AD traits might be particularly helpful for improving model performance [34, 48]. Finally, leveraging the rapid advances in machine learning suggests that it may be possible to impute and include underlying AD pathologies to improve model performances [23, 49-53].

### Strength & Limitations

The study has several strengths that lend confidence for the current findings. All subjects were recruited from the community, underwent an annual detailed clinical evaluation, and were determined to be free of dementia at study entry. Large numbers of men and women underwent annual assessments, and follow-up rates were very high. Uniform, structured procedures were followed that enhanced stability of diagnoses across time, space, and examiners. Diverse non-cognitive covariates were examined. An important strength of the current study design is that we compared the performances of models predicting cognitive impairment and Alzheimer’s dementia in the same individuals with identical covariates [54]. Moreover, we were able to replicate our results in an independent sample of older adults who underwent the same assessments and testing by the same staff. However, further studies will be needed due to limitations. Participants were predominantly Americans of European descent and have higher than average levels of education, so our findings will need to be replicated in more diverse populations. Brain imaging and fluid biomarkers were not examined. While we employed the NIA-AA criterion for clinical diagnosis of Alzheimer’s dementia, we recognize that this diagnosis is often associated with mixed brain pathologies as shown in our analysis of decedents who underwent brain autopsy. The particular cognitive battery employed is more extensive than what might be available outside of a research setting highlighting the importance of our findings using the MMSE.

## Supporting information

Suppemental data

## Acknowledgments

This work was supported by the National Institute of Health R35GM138313, P30AG10161, K01AG054700, R01AG15819, R01AG17917, R01AG56352; the Illinois Department of Public Health; and the Robert C. Borwell Endowment Fund. The funding organizations had no role in the design or conduct of the study; collection, management, analysis, or interpretation of the data; or preparation, review, or approval of the manuscript. We are deeply indebted to all participants who contributed their data and agreed to autopsy at the time of their death. We are thankful to the staff at the Rush Alzheimer’s Disease Center.

