## Supplementary material for "Risk models based on non-cognitive measures may identify presymptomatic Alzheimer’s disease": Suppemental data

### Supplementary Figures

**Supplementary Figure 1. Duration of follow-up in adults with no dementia (NCI or MCI) at baseline (A) or adults with normal cognition (NCI) at baseline (B).**

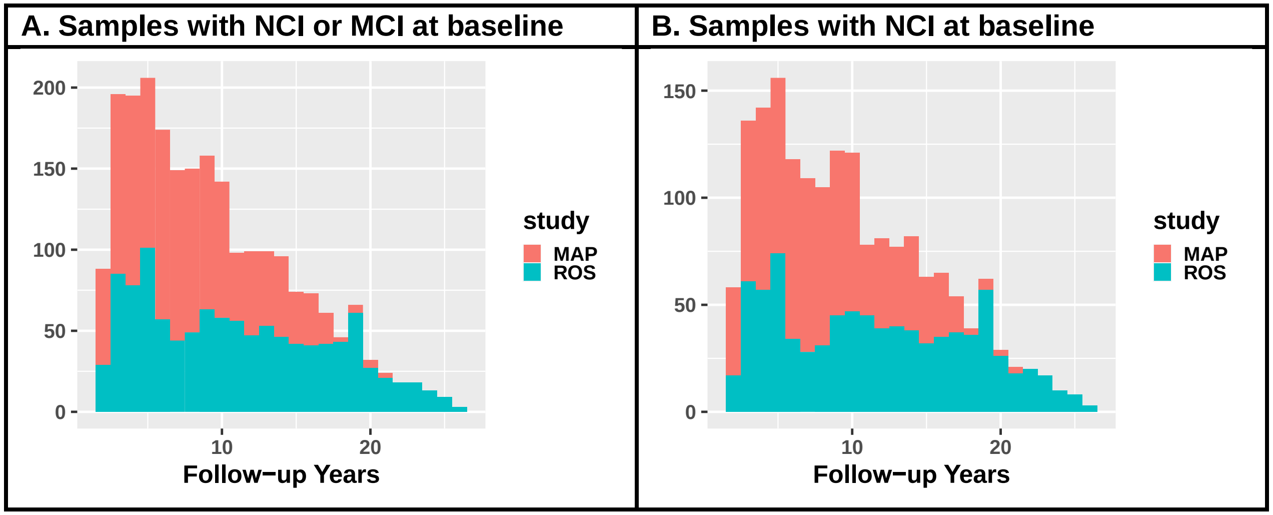

**Supplementary Figure 2. Correlation heatmap of all clinical covariates considered in the Cox models.**

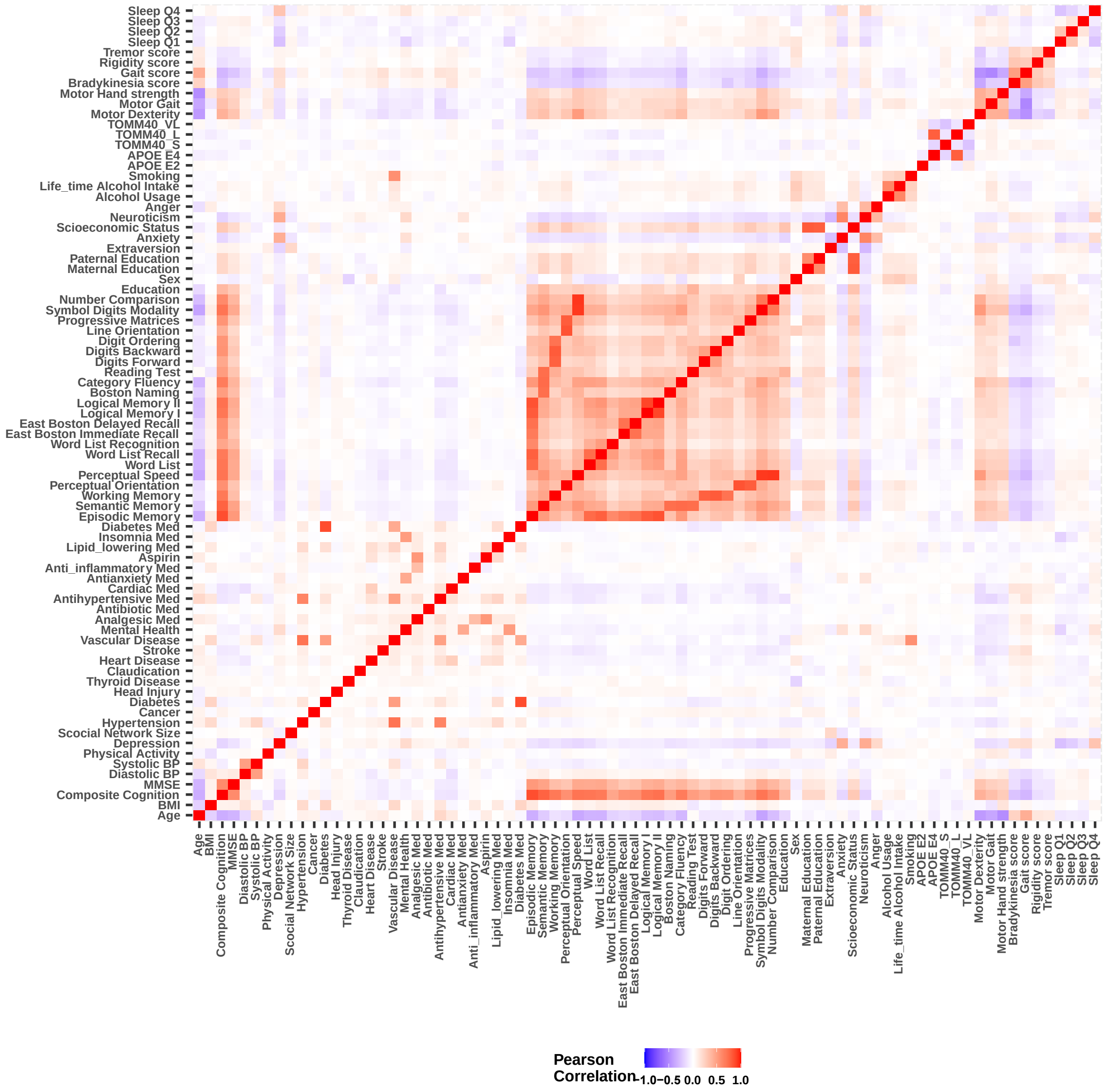

**Supplementary Figure 3. ROC plots of risk prediction for incident Alzheimer's dementia and incident cognitive impairment with different analytic cohorts and combinations of cognitive covariates as shown.** Model D uses composite cognition alone; Model E uses MMSE alone; Model F uses 5 cognitive ability scores; Model G uses 17 individual cognitive test scores.

**ROW 1 No dementia (NCI or MCI) at baseline and incident Alzheimer's Dementia:** Models D, F, G have comparable prediction performance with p-values>0.176 for Year (3, 5), which all significantly better than Model E with p-values=~($1.72 \times 10^{-6}, \ 8.57 \times 10^{-7}$) for Year (3, 5). Model E performed comparable with Model A with only non-cognitive covariates in Fig 1 with p-values = (0.313, 0.112) for Year (3, 5).

**ROW 2 NCI at baseline and incident Alzheimer's Dementia:** Models D, F, G have comparable prediction performance, which all significantly better than Model E with p-values=~(0.05, 0.04) for Year (3, 5). Model E has comparable performance with Model A with only non-cognitive covariates in Fig 1 for Year 3 with p-value=0.45, but significant poorer performance in Year 5 with p-values=0.012.

**ROW 3 NCI and incident cognitive impairment:** Models D, F, G have comparable prediction performance with p-values > 0.45, which all significantly better than Model E. Model E performed significant poorer than Model A with only non-cognitive covariates in Fig 1 with p-values=(0.005, 0.001) for Year (3, 5).

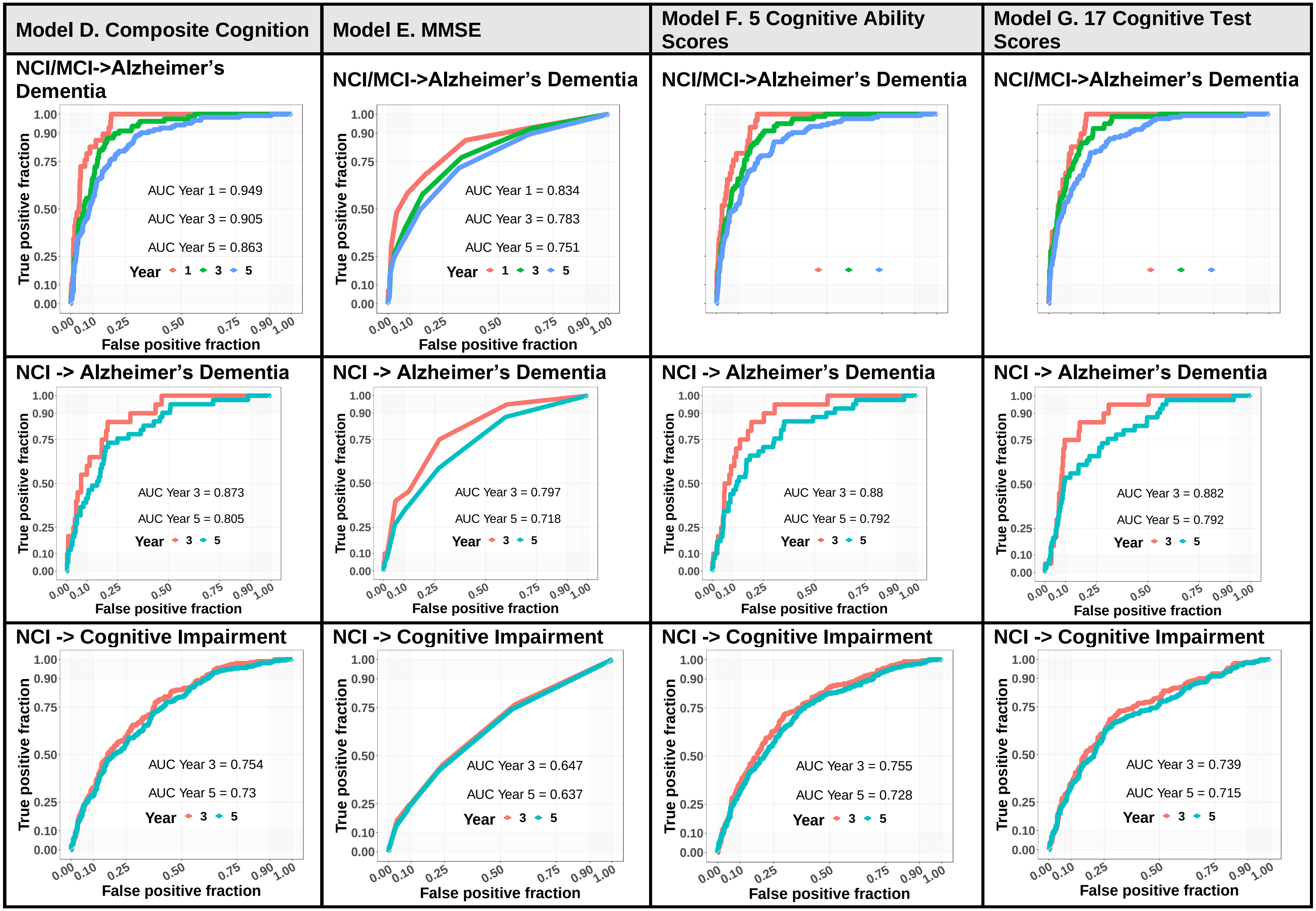

Model D. Composite Cognition
Model E. MMSE
Model F. 5 Cognitive Ability Scores
Model G. 17 Cognitive Test Scores
NCI/MCI->Alzheimer's Dementia
AUC Year 1 = 0.949
AUC Year 3 = 0.905
AUC Year 5 = 0.863
Year 1 3 5
NCI/MCI->Alzheimer's Dementia
AUC Year 1 = 0.834
AUC Year 3 = 0.783
AUC Year 5 = 0.751
Year 1 3 5
NCI/MCI->Alzheimer's Dementia
NCI/MCI->Alzheimer's Dementia
NCI -> Alzheimer's Dementia
AUC Year 3 = 0.873
AUC Year 5 = 0.805
Year 3 5
NCI -> Alzheimer's Dementia
AUC Year 3 = 0.797
AUC Year 5 = 0.718
Year 3 5
NCI -> Alzheimer's Dementia
AUC Year 3 = 0.88
AUC Year 5 = 0.792
Year 3 5
NCI -> Alzheimer's Dementia
AUC Year 3 = 0.882
AUC Year 5 = 0.792
Year 3 5
NCI -> Cognitive Impairment
AUC Year 3 = 0.754
AUC Year 5 = 0.73
Year 3 5
NCI -> Cognitive Impairment
AUC Year 3 = 0.647
AUC Year 5 = 0.637
Year 3 5
NCI -> Cognitive Impairment
AUC Year 3 = 0.755
AUC Year 5 = 0.728
Year 3 5
NCI -> Cognitive Impairment
AUC Year 3 = 0.739
AUC Year 5 = 0.715
Year 3 5
True positive fraction
False positive fraction
0.00 0.10 0.25 0.50 0.75 0.90 1.00

**Supplementary Figure 4. Selected predictive covariates risk prediction for incident Alzheimer's dementia and incident cognitive impairment with different analytic cohorts and combinations of cognitive covariates as shown.**
**Row 1 Covariates of 5 cognitive ability scores.** Episodic memory is the top protective factor for both cognitive outcomes.
**Row 2 Covariates of 17 individual cognitive test scores.** East Boston delayed recall test, Boston naming, and word list recall tests are leading protective factors for both cognitive outcomes.

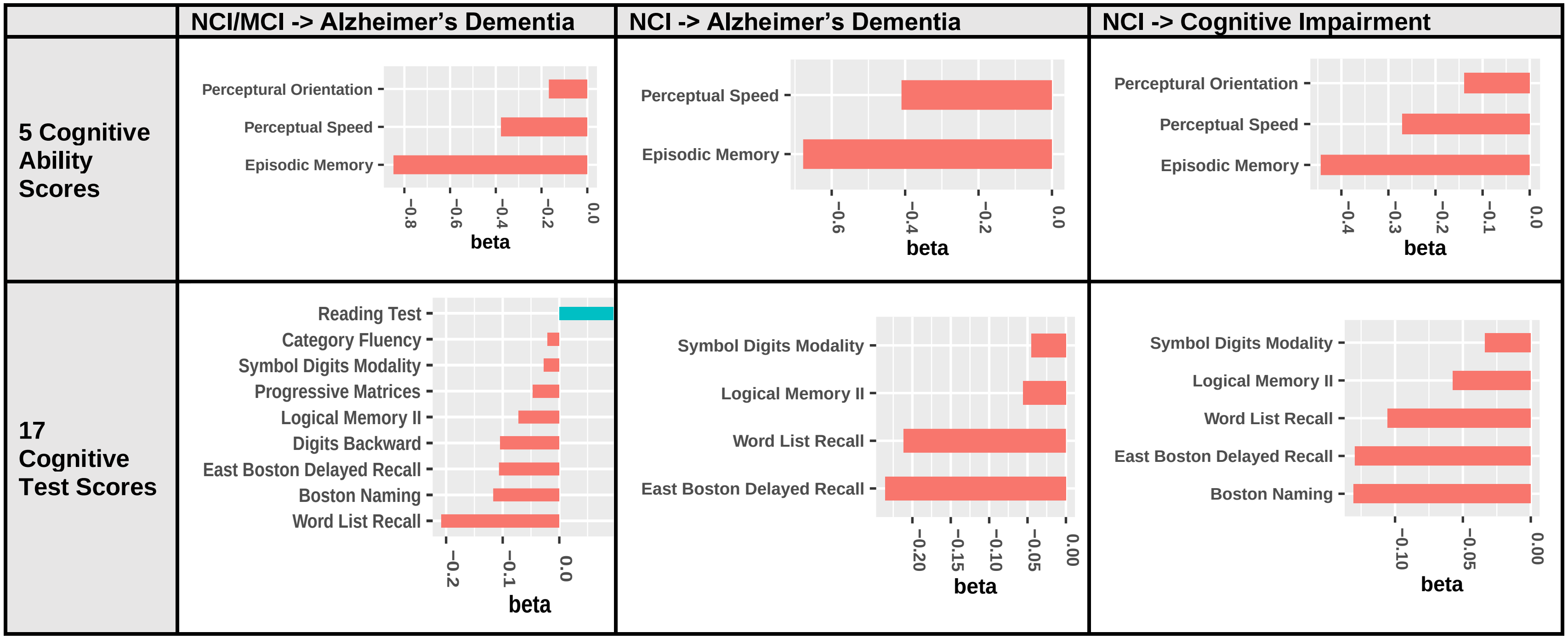

**Supplemental Figure 5. Association between predicted risk scores of developing Alzheimer's dementia in 3, 5 years and 12 ADRD neurodegenerative and cerebrovascular disease brain pathologies.** Test risk scores were generated for the test ROS cohort by the full Model C as in Figure 1, Column 3, Row 1. A total of 12 ADRD neurodegenerative and cerebrovascular disease brain pathologies were tested — Amyloid, Tangles, Global AD pathology (gpath), NIA Reagan pathology (NIA), Lewy body pathology (LewyBody), Hippocampal Sclerosis (Hippo), TDP-43, Macroinfarct (Macro), Microinfarct (Micro), Atherosclerosis (Arthero), Arteriolosclerosis (Arteriol), Cerebral amyloid angiopathy (CAA). The test $-log10(Pvalues)$ were plotted and the significance threshold with Bonferroni correction for testing 12 pathologies ($-log10(0.05/12)$) was plotted as the blue line. A total of 5 out of 12 pathologies were associated with the risk model (above the blue line), including neurodegenerative pathology (Tangles, Global AD pathology, NIA Reagan) and cerebrovascular disease pathologies (Atherosclerosis and Cerebral amyloid angiopathy).

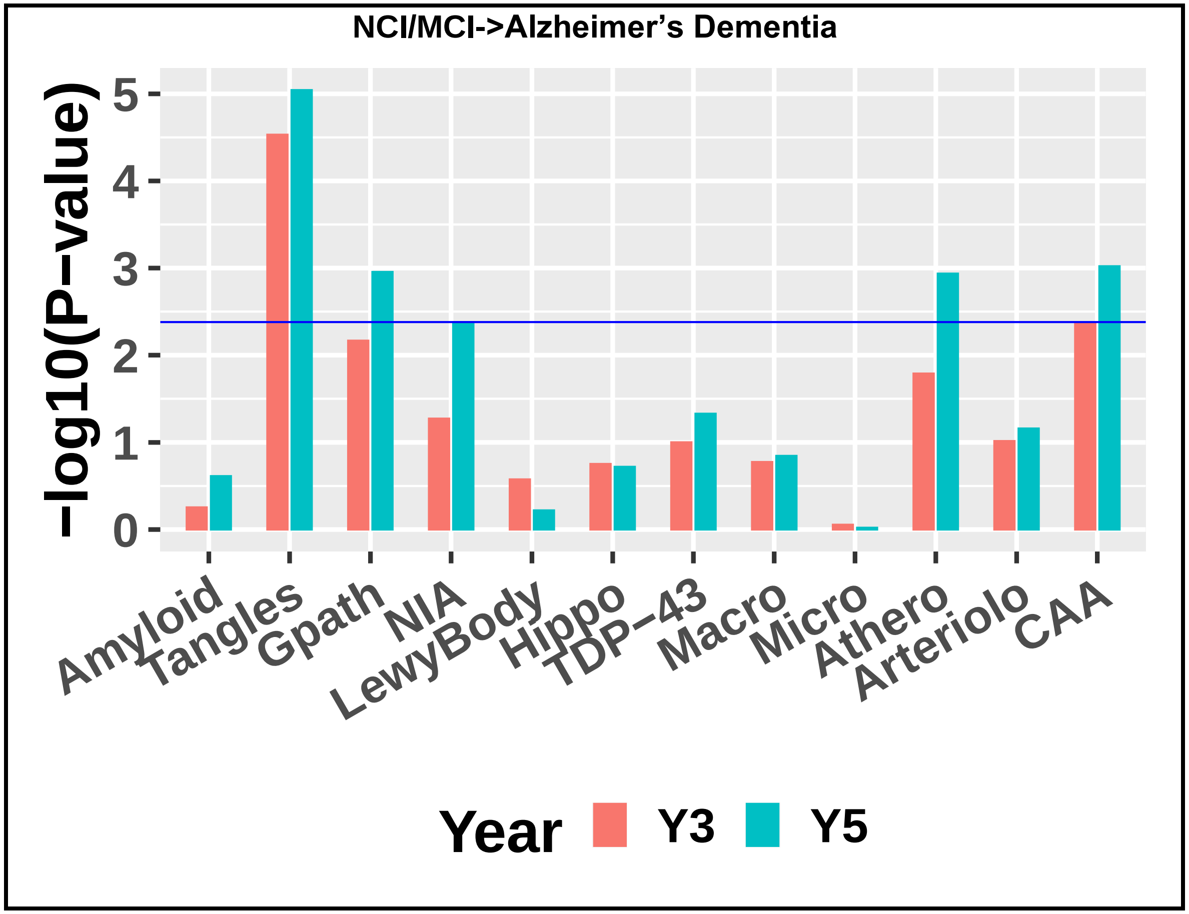

#### Supplemental Tables

##### Supplementary Table 1: Sample sizes per event type for studying incident Alzheimer’s dementia.

| | NCI or MCI (Right Censored) | Alzheimer’s Dementia | Death | Row Total |
|---|---|---|---|---|
| **Baseline NCI** | 976 | 369 | 397 | 1742 |
| **Baseline MCI** | 195 | 245 | 100 | 540 |
| **Column Total** | 1171 | 614 | 497 | 2282 |

##### Supplementary Table 2: Sample sizes per event type for studying incident cognitive impairment.

| | NCI (Right Censored) | Cognitive Impairment (MCI or Alzheimer’s Dementia) | Death | Row Total |
|---|---|---|---|---|
| **Baseline NCI** | 643 | 957 | 176 | 1776 |

**Supplementary Table 3. Additional clinical variables of the analytic cohort at baseline (n=1179 in MAP; n=1103 in ROS).**

| Group | Variable | Mean (SD) or N (%) |
|---|---|---|
| Clinical and Chronic Health Variables | Diastolic blood pressure (40, 122.5) | 74.47 (11.90) |
| | Hypertension blood pressure (83, 215.5) | 134.13 (18.29) |
| | Depression score (0, 9) | 0.94 (1.48) |
| | Hypertension (N) | 1142 (49.93) |
| | Cancer (N) | 719 (31.43) |
| | Diabetes (N) | 277 (12.11) |
| | Head injury (N) | 148 (6.47) |
| | Thyroid disease (N) | 440 (19.23) |
| | Claudication (N) | 138 (6.03) |
| | Heart disease (N) | 208 (9.09) |
| | Stroke (N) | 156 (6.82) |
| | Cardiovascular disease history counts (0, 3) | 0.93 (0.78) |
| | Alcohol usage in the past year, grams per day (0, 116.55) | 4.56 (11.14) |
| | Alcohol usage when drank most in lifetime (0, 6) | 0.46 (0.98) |
| | Smoking (never smoked 0, former smoker 1, current smoker 2) | 0.33 (0.51) |
| Medication Usage | Mental health (N) | 529 (23.13) |
| | Analgesic (N) | 1677 (73.32) |
| | Antibiotic (N) | 158 (6.90) |
| | Anti-hypertensive (N) | 1394 (60.95) |
| | Cardiac (N) | 228 (9.96) |
| | Anti-anxiety (N) | 133 (5.81) |
| | Anti-inflammatory (N) | 561 (24.52) |
| | Aspirin (N) | 992 (43.37) |
| | Lipid lowering (N) | 740 (32.35) |
| | Insomnia (N) | 171 (7.47) |
| | Diabetes (N) | 203 (8.87) |

**Supplementary Table 4. Five groups of covariates considered by Cox proportional hazard models.**

| Group | Variables |
|---|---|
| **Demographics and Common Alzheimer's Risk Factors** | Age, Sex, Education, MMSE Score, APOE Genotype E2 and E4 |
| **Clinical and Chronic Health Variables** | BMI, Blood pressure (systolic, diastolic), Depression score, Hypertension, Cancer, Diabetes mellitus, Head injury, Thyroid disease, Claudication, Heart disease, Stroke, Cardiovascular disease, Alcohol usage (in the past year and when drank most in lifetime), Smoking, |
| **Medication Usage** | Usage of mental health, Analgesic, Antibiotic, Anti-hypertensive, Cardiac, Anti-anxiety, Anti-inflammatory, Aspirin, Lipid lowering, Insomnia, Diabetes medications |
| **Covariates Uniquely Profiled by ROS/MAP** | Composite cognition test score, Physical activity, Social network size, Maternal and paternal education, Anxiety, Neuroticism indicating proneness to psychological distress, Early life socioeconomic status, Anger trait, TOMM40 haplotype (S, L, VL) |
| **Motor and Sleep Metrics** | Dexterity, Gait function, Hand strength, Bradykinesia, Parkinsonian gait, Rigidity, Tremor; self-reported sleep latency, sleep consolidation, daytime sleepiness, sleep quality |

**Supplementary Table 5. Top 5 significant selected predictive covariates in the Cox models considering non-cognitive and MMSE covariates (Model B in Figs 1-2).** Estimated log hazard ratio (Beta) and the corresponding standard deviation (Beta SD), as well as p values are listed. All covariates were sorted by their p-values.

| | Covariates | Beta (Log Hazard Ratio) | Beta SD | P value |
|---|---|---|---|---|
| **NCI/MCI -> Alzheimer's Dementia** | MMSE | -0.413 | 0.048 | $6.36 \times 10^{-18}$ |
| | Age | 0.613 | 0.087 | $1.60 \times 10^{-12}$ |
| | *APOE* E4 | 0.291 | 0.054 | $6.90 \times 10^{-8}$ |
| | Hand strength | -0.316 | 0.077 | $3.97 \times 10^{-5}$ |
| | APOE E2 | -0.227 | 0.072 | $1.62 \times 10^{-3}$ |
| **NCI -> Alzheimer's Dementia** | Age | 0.814 | 0.069 | $4.26 \times 10^{-14}$ |
| | *APOE* E4 | 0.344 | 0.054 | $6.39 \times 10^{-7}$ |
| | Motor Gait | -3.096 | 0.086 | $3.33 \times 10^{-4}$ |
| | MMSE | -0.206 | 0.059 | $4.82 \times 10^{-4}$ |
| | Hand strength | -0.274 | 0.099 | $5.90 \times 10^{-3}$ |
| **NCI -> Cognitive Impairment** | Age | 0.402 | 0.061 | $4.59 \times 10^{-11}$ |
| | MMSE | -0.243 | 0.046 | $1.66 \times 10^{-7}$ |
| | *APOE* E4 | 0.204 | 0.045 | $5.82 \times 10^{-6}$ |
| | Parkinsonian gait score | 0.185 | 0.046 | $6.60 \times 10^{-5}$ |
| | Daytime sleepiness | 0.161 | 0.052 | $2.15 \times 10^{-3}$ |

**Supplementary Table 6. Recent risk prediction models of Alzheimer's dementia.** The C-statistic mentioned in this table is equivalent to the AUC used in our study.

| No Cognitive Variable Examined | | | | |
|---|---|---|---|---|
| **Study** | **Demographics** | **Examined Variables** | **Final Variables in the Models** | **Model Discrimination and Follow-up** |
| Kivipelto M, et al. PMID= 16914401 | N=1409; F=875 (62%); age (mean=50.4; SD=6.0; min=39; max=64) | systolic blood pressure (SBP), diastolic blood pressure (DBP), BMI, total cholesterol, smoking, physical activity, age, sex, education | Age, sex, education, SBP, BMI, physical activity, total cholesterol | Follow up (mean=20.9; SD=4.9); outcome=dementia(n=61); AUC=0.77 (95%CI: 0.71 – 0.83). |
| Li J, et al. PMID= 28627378 | N=2383; age (min=60; max=88; 55% (n=1349) were less than 70); F=1441 (60%) | BMI, smoking, alcohol, coffee or tea, low salt diet, comorbidity (hypertension, DM, stroke, cancer, vascular diseases of the brain [embolic infarct, ICH, SAH, other vascular diseases), ischemic attack (possibly TIA) | Age, marital status, BMI, stroke, DM, cancer, ischemic attack | C=0.716; follow up: max=30; Outcome=dementia, n=778; |
| Walters K et al. PMID= 26797096 | Has development and validation datasets. Developed dementia risk models for two age groups: 60-79, 80-95.<br>Age 60-79 [development dataset (N=800013) and validation (N=226140). Age (mean=65.6; SD=6.1); F=51.8%;].<br>Age 80-95 [development dataset (N=130382) and validation (N=38084). Age (mean=84.8; SD=3.9); F=66%]. | total cholesterol, HDL, weight, SBP, height, BMI, local area deprivation score, marital status, history of (stroke, cancer, alcohol, smoking, DM, coronary heart disease, AF), drugs (antidepressant, anxiolytics, hypnotics, antihypertensive, statin, NSAIDs, aspirin, age, sex) | 60-79: age, sex, calendar year at baseline, BMI, smoking, history of (stroke, AF, DM, alcohol), drug (antidepressant, aspirin).<br>80-95: age, sex, calendar year at baseline, BMI, antihypertensive drug, SBP, lipid ratio, smoking, history of (stroke, AF, DM, alcohol), drug (antidepressant, anxiolytics, NSAIDs, aspirin). | 60-79 (validation dataset): C=0.84 (95% CI: 0.81 – 0.87); dementia, n=1923; FU=median of 5 years;<br><br>80-95 (validation dataset): C=0.56 (95% CI: 0.55 – 0.58); dementia, n=1699; FU=median of 3.9 years; |
| Reitz C, et al. PMID= 20625090 | N=1051; age=75.7 (6.3); F=696 (66.2%); | Age, sex, education, ethnicity, APOE E4, DM, hypertension, heart disease, smoking, HDL, waist to hip (WHP); | Age, sex, education, ethnicity, APOE E4, DM, hypertension, heart disease, smoking, HDL, waist to hip (WHP); | No discrimination is provided.<br>Outcome=AD dementia (n=92); FU (mean=4.0; SD=1.4) |
| Middelaar TV, et al. PMID= 29960597 | N=3339; age (range:70-79; mean=74; SD=2.5); F=1492 (44.6%); | Libra index: Age, education, depression, hypertension, obesity, smoking, cholesterol, DM, renal dysfunction, physical inactivity, coronary heart disease, alcohol | | Outcome=dementia(n=220); Libra index was not associated with dementia. |
| Anstey KJ, et al. PMID: 24465922 | Three cohorts the risk score was examined; | Age, sex, DM, Traumatic brain injury, Cognitive activity, Social | The Australian National University AD Risk Index | MAP: [(FU: mean=3.5, SD=3.0); 17.5% developed |

| | | | | |
|---|---|---|---|---|
| | Rush Memory and Aging Project (MAP, N=903); Age (mean=79.8, SD=7.4); F=75.2%;<br>The Kungsholmen Project (KP, N=905); Age (mean=81.5, SD=5.0); F=75.0%;<br>The Cardiovascular Health Cognition Study (CVHS, N=2496); Age (mean=72.3, SD=4.9); F=59.1%; | network and engagement, Smoking, Alcohol, Physical activity, Fish intake, Depression symptoms | (ANU-ADRI); includes all the mentioned variables. | AD]; C=0.733 (0.691 – 0.776)<br>KP: [(FU: mean=3.5, SD=3.0); 20.0% developed AD]; C=0.637 (0.596 – 0.678)<br>MAP: [(FU: mean=3.5, SD=3.0); 11.1% developed AD]; C=0.740 (0.712 – 0.768) |
| Licher S et al.<br>PMID: 29740780 | N=6667; Age= (mean=69.1, SD=8.2); F=56.8%; | Four dementia risk score examined: CAIDE, ANU-ADRI, Brief Dementia Screening Indicator (BDSI), Dementia Risk Score (DRS). | BDSI: age, education, BMI, history (DM, stroke), assistance needed with finances and medications, depressive symptoms.<br>DRS: Age, sex, BMI, history (hypertension, alcohol, smoking, DM, stroke, atrial fibrillation), depressive symptoms, social deprivation, anxiety, aspirin, NSAIDS, calendar year | FU=(median=13.2, Q1=10.1, Q3=16.3); outcome: dementia(n=867);<br>The 10-year discrimination:<br>CAIDE: C=0.55 (0.53-0.58);<br>ANU-ADRI: C=0.75 (0.74-0.77);<br>BDSI: C=0.78 (0.76-0.81);<br>DRS: C=0.81 (0.78-0.83);<br>The authors examined age alone and the C statistics did not change (except CAIDE that did not have age component).<br>The 10-year discrimination examining age component alone:<br>ANU-ADRI: C=0.77 (0.75-0.79);<br>BDSI: C=0.81 (0.78-0.83);<br>DRS: C=0.81 (0.78-0.83);<br>The 10-year discrimination examining components other than age:<br>ANU-ADRI: C=0.52 (0.49-0.54);<br>BDSI: C=0.60 (0.58-0.63);<br>DRS: C=0.57 (0.55-0.60); |

**Cognitive Variable Examined**

| Study | Demographics | Examined Variables | Final Variables in the Models | Model Discrimination and Follow-up |
|---|---|---|---|---|

| | | | | |
|---|---|---|---|---|
| Lin H, et al. PMID: 25788555 | N=72127; F=56.0%; Age=70-74 (50%); | Age, sex, BMI, Hypertension, FBS, A1c, Kihon frailty index items (IADL, physical function, nutritional status, oral function, house bandedness, subjective cognitive function, risk of depression) | 3 models examined;<br>Model1: age-sex<br>Model2: Model1+Frailty index (that includes subjective cognitive function)<br>Model3: Model2+health factors (FBS, BMI) | C:<br>Model1: 0.734 (0.728–0.741<br>Model2: .791 (0.785–0.797)<br>Model3: 0.788 (0.782–0.794)<br>FU (mean=3.3; max=4.2 years); Outcome=dementia; |
| Steenland K, et al. PMID: 29843232 | N=424; Age (mean=72.6, SD=7.1); Female (n=181, 42.7%); participants were amnestic MCI. | Age. Gender, education, race, whole brain volume, medial temporal volume, hippocampal volume, CSF measures of Aβ and tau, APOE E4, MMSE, ADAS-Cog, and summary measures for memory (derived from ADAS-Cog, Rey Auditory verbal Learning Test, Logical Memory, MMSE) and executive function tests (derived from category fluency, Trails A and B, Digit span backward, Digit-Symbol test, Clock drawing test. Also, Functional Activity Questionnaire and *APOE* ε4 were examined. | Summary memory measure, APOE E4, tau/ Aβ ratio, hippocampal volume, Functional activity questionnaire, | FU (median=4, Q1=3, Q3=5) years;<br>Outcome=AD dementia n=150;<br>C=0.91.<br>This paper also included in separate cohort NCI participants with the outcome of amnestic MCI. It is below (C=0.80). |
| Rosenberg A, et al. PMID: 31805990 | From MCI; Age (mean=64.8, SD=9.1); Female (n=174, 54.7%); | | Basic model: age, Rey Auditory Verbal Learning Test;<br>CSF model: Basic model+ Aβ+tau<br>APOE E4model: Basic+ APOE E4<br>SBP Model: Basic + SBP<br>BMI Model: Basic + BMI<br>Depressive model: Basic + Depressive symptoms | Outcome: dementia (n=121);<br>FU=(mean=2.8, SD=1.9, range: 1-10);<br>Basic Model: C=0.69<br>CSF Model: C=0.78<br>APOE E4 Model: C=0.72<br>SBP Model: 0.70<br>BMI Model: 0.74<br>Depressive model: 0.70 |
| Zandifar A, et al. PMID: 31931400 | From amnestic MCI; N=756; Age (mean=73, SD=7); F=40% | SNIPE MRI measures (in hippocamp and entorhinal cortex), which is a computed voxel-based index to differentiate AD dementia from normal control; neurocog measures (ADAS-cog, Rey Auditory Verbal Learning Test, MMSE). The values corrected for age, sex, education. | Combined model performed better than single predictor. In single predictors: ADAS-cog>hippocampus SNIPE>MMSE~RAVLT~Entorhinal SNIPE>age~sex~educ | FU=up to 108 months.<br>Modeling was done for different FU: 12, 24, 36, ..., 108.<br>AUC=0.85 – 0.92 for f=FU from 24 to 84 months.<br>Outcome: AD dementia ( |

| | | | | |
|---|---|---|---|---|
| Ewers M, et al. PMID: 21159408 | From MCI and HC subjects (N=312); MCI subjects who had not progressed were excluded. | Hippocampal volume, entorhinal cortex thickness, CSF tau and Aβ, age, sex, *APOE* ε4, neuropsych (RAVLT, Digit Symbol substitution test, TMT-A and B, category fluency, digit span forward and backward, Boston naming test) | Age, sex, and *APOE* ε4 were not associated with the outcome in the presence of CSF and MRI measures. The best model included RAVLT, TMT-B, and CSF t-tau/ Aβ. | FU (mean=2.3, SD=0.6, Max=3.3); Outcome: AD dementia (n=81) The best model discriminating AD from HC had classification accuracy of 94.8%. |
| Barnes, DE, et al. PMID: 24495339 | From amnestic MCI (N=382); age (mean=75, SD=7); F=(n=137, 36%); | Age, sex, race, education, marital status, family history of AD, premorbid IQ, medical history (stroke, other vascular diseases, depression, hypertension, diabetes, smoking, respiratory condition, cancer, kidney disease, head injury, thyroid disease), symptoms and vital signs (low energy, insomnia, abnormal gait, SBP, DBP, pulse rate, BMI, Functional Assessment Questionnaire, Neuropsychiatric inventory, depressive symptoms), MRI (including hippocampal volume and entorhinal cortex thickness), blood (*APOE* ε4, Aβ40), cognitive tests (including RAVLT, TMT a & B, …) | FAQ, middle temporal cortex thickness, hippocampal subcortical volume, ADAS-Cog, Clock test. | Outcome=AD dementia (n=179); FU (Max: 4.0); C=0.78 (0.75 – 0.81) |

**Supplementary Table 7. Recent risk prediction models of cognitive impairment (MCI or Alzheimer's dementia).**

| Study | Demographics | Examined Variables | Final Variables in the Models | Model Discrimination and Follow-up |
|---|---|---|---|---|
| **No Cognitive Variable Examined** | | | | |
| Fouladvand F, et al. PMID: 33194303 | N= 3,265; I did not find any data about the demographic info | Age, sex, education, history (HTN, DM, AF, angina, CABG, MI, CHF), neuropsychiatric symptoms, activity of daily living, slow gait, cognitive complaint, memory concern | It included all the variables; However, random forest indicated the top most contributing variables were: age, hypertension, education, depression, anxiety | MCI (n=558); No AUC or C was reported. The prediction accuracy was 0.82 and precision was 0.44. |
| **Cognitive Variable Examined** | | | | |
| Pankratz VS, et al. PMID: 25788555 | N=1449; F=50%; Age≥80: 41%; | Education, subjective memory complaint, alcohol, stroke, DM, atrial fibrillation, smoker, midlife dyslipidemia, hypertension, BMI≥30, marital status, presence of agitation, apathy, anxiety, Clinical Dementia Rating (CDR), *APOE* ε4, short test of mental status, Unified Parkinson's Disease Rating Scale score, Functional Activities Questionnaire, slow gait, Depression | They developed several models. Basic model including demographics and clinical history variables. Augmented model without *APOE* ε4: including cognition score, CDR, motor scores, neuropsychiatric scores. Augmented model with *APOE* ε4 | FU (median=4.8, Q1=2.5, Q3=6.4) MCI=27.7% (n=401); Basic model: C=0.60 (SE=0.3); Augmented model without *APOE* ε4: C=0.70 (0.03); Augmented model with *APOE* ε4: C=0.70 (0.03); These C statistics were cross-validated ones. The originals were the same or 0.02 above. |
| Steenland K, et al. PMID: 29843232 | N=224; Age (mean=74.4, SD=5.8); Female (n=109, 48.7%); | Age. Gender, education, race, whole brain volume, medial temporal volume, hippocampal volume, CSF measures of Aβ and tau, Apoe ε4, MMSE, ADAS-Cog, and summary measures for memory (derived from ADAS-Cog, Rey Auditory verba Learning Test, Logical Memory, MMSE) and executive function tests (derived from category fluency, Trails A and B, Digit span backward, Digit-Symbol test, Clock drawing test. Also, Functional Activity Questionnaire and *APOE* ε4 were examined. | Summary memory measure, tau/ Aβ ratio, hippocampal volume, | FU (median=4, Q1=3, Q3=5)years; Outcome=Amnestic MCI, n=37; C=0.80. This paper also included in separate cohort aMCI participants with the outcome of AD dementia. It is above (C=0.91). |
| Albert M, et al. PMID: 29365053 | N=224; Age (mean=56.9, SD=8.4), Female=62.1% | Memory test and digit symbol substitution test, Aβ, tau from CSF, right hippocampal volume, | Full model includes all the variables; Efficient model includes every variable except CSF Aβ; | Follow up till 10 years, AUC calculated at 5, 7, and 10 years. |

| | | | | |
|---|---|---|---|---|
| | | right entorhinal cortex thickness, *APOE* ε4, age, education, | Demographic model includes only age and education. | Outcome: MCI or AD dementia, n=46;<br>Full model, 5Y=0.850 (0.807 – 0.913)<br>Full model, 10Y=0.831 (0.781 – 0.890)<br>Efficient model, 5Y=0.849 (0.802 – 0.910)<br>Efficient model, 10Y=0.822 (0.769 – 0.886)<br>Demog model, 5Y=0.681 (0.614 – 0.770)<br>Demog model, 10Y=0.680 (0.612 – 0.756) |
